# Genetic epidemiology of SARS-CoV-2 transmission in renal dialysis units - a high risk community-hospital interface

**DOI:** 10.1101/2021.03.24.21253587

**Authors:** Kathy K Li, Y. Mun Woo, Oliver Stirrup, Joseph Hughes, Antonia Ho, Ana Da Silva Filipe, Natasha Johnson, Katherine Smollett, Daniel Mair, Stephen Carmichael, Lily Tong, Jenna Nichols, Elihu Aranday-Cortes, Kirstyn Brunker, Yasmin A. Parr, Kyriaki Nomikou, Sarah E McDonald, Marc Niebel, Patawee Asamaphan, Vattipally B Sreenu, David L Robertson, Aislynn Taggart, Natasha Jesudason, Rajiv Shah, James Shepherd, Josh Singer, Alison H.M. Taylor, Zoe Cousland, Jonathan Price, Jennifer S. Lees, Timothy P.W. Jones, Carlos Varon Lopez, Alasdair MacLean, Igor Starinskij, Rory Gunson, Scott T.W. Morris, Peter C. Thomson, Colin C. Geddes, Jamie P. Traynor, Judith Breuer, The COVID-19 Genomics UK (COG-UK) consortium, Emma C. Thomson, Patrick B. Mark

## Abstract

**Objectives:** Patients requiring haemodialysis are at increased risk of serious illness with SARS-CoV-2 infection. To improve the understanding of transmission risks in six Scottish renal dialysis units, we utilised the rapid whole-genome sequencing data generated by the COG-UK consortium.

**Methods:** We combined geographical, temporal and genomic sequence data from the community and hospital to estimate the probability of infection originating from within the dialysis unit, the hospital or the community using Bayesian statistical modelling and compared these results to the details of epidemiological investigations.

**Results:** Of 671 patients, 60 (8.9%) became infected with SARS-CoV-2, of whom 16 (27%) died. Within-unit and community transmission were both evident and an instance of transmission from the wider hospital setting was also demonstrated.

**Conclusions:** Near-real-time SARS-CoV-2 sequencing data can facilitate tailored infection prevention and control measures, which can be targeted at reducing risk in these settings.

## Introduction

The emergent SARS-CoV-2 virus which causes COVID-19 is associated with increased morbidity and mortality in older individuals and in those with chronic diseases.^1,2^ Chronic kidney disease and pre-existing conditions that may increase the risk of renal failure, such as cardiovascular disease, diabetes mellitus and hypertension, are significantly associated with an increased risk of death in COVID-19.^3,4^ Individuals requiring haemodialysis in hospital are also at increased risk of nosocomial infection due to prolonged outpatient dialysis (typically three times weekly for four hours per session) and to community infection due to regular travel on public or hospital transport, often with other patients.^5-7^ The case fatality rate in dialysis patients has been reported as 20-30% compared with 1-2% in patients not requiring haemodialysis.^8-11^

Whole-genome pathogen sequencing has become increasingly accessible. Its utility in the context of evolving outbreaks has been demonstrated with Ebola, Zika and hospital outbreak investigations.^12-15^ The COVID-19 Genomics UK (COG-UK) Consortium, funded by the UK Department of Health and Social Care, UK Research and Innovation (UKRI) and the Wellcome Sanger Institute, was set up to enable real-time sequencing at a population level and to facilitate the investigation and management of hospital-associated infections, providing policymakers with information on introductions and transmission events.^16-18^

We aimed to investigate the genetic epidemiology of COVID-19 infection from patients attending six Scottish renal dialysis unit(s) (RDU) using a Bayesian statistical analysis framework incorporating temporal, geographical and genetic sequence data. These results were evaluated alongside traditional epidemiological investigations. We further investigate the clinical impact of COVID-19 infection on haemodialysis patients and incorporate the information obtained using combined genetic and epidemiological data to inform future infection control strategies.

## Methods

### Study design and participants

The Glasgow Renal and Transplant Unit based at the Queen Elizabeth University Hospital and University Hospital Monklands serve a combined population of approximately 2.16 million people across West and Central Scotland under NHS Greater Glasgow and Clyde, NHS Forth Valley and NHS Lanarkshire Health Boards. These institutions provide haemodialysis treatment for 828 outpatients across eight RDUs (numbers extracted 1^st^ March 2020). Use of anonymised data was approved by the Local Privacy Advisory Group of NHS Greater Glasgow and Clyde ‘Safe Haven’ on behalf of the West of Scotland Ethics Committee (approval GSH/20/RE/001). Follow up was until 4^th^ June 2020. From 2^nd^ March 2020, patients attending for dialysis with symptoms of COVID-19 were tested for SARS-CoV-2 by nasopharyngeal swab. We report data on the six RDUs (RDU 1-6, number of patients treated with dialysis =671) with any patients with SARS-CoV-2 infection. Initially, personal protective equipment (fluid-resistant surgical masks, eye protection, aprons and gloves) (PPE) was *not* recommended by UK and Scottish Governments for HCWs caring for patients, unless clinical index of suspicion was high for COVID-19 or for aerosol generating procedures. RDU1 instigated PPE for HCWs and surgical masks for patients whilst travelling to, from and during dialysis on 23^rd^ March in response to earlier infections. From 3^rd^ April 2020, the UK-recommendations changed to include any close patient contact. Surgical masks were also given to all patients, as per RDU1 and shared-patient transport discontinued. Until 5^th^ April HCWs were ineligible for SARS-CoV-2 testing unless hospitalised, being advised to self-isolate for 7 days in keeping with Scottish government policy.

### Laboratory diagnosis

Nasopharyngeal swabs in viral PCR solution (1:1 ratio of EasyMag Nuclisens Extraction Buffer (BioMerieux, France) and EMEM) were extracted and tested according to the availability of assays at the diagnostic laboratory.^i^ Surplus RNA extract was collected for sequencing with ethical approval from the NHSGGC biorepository (16/WS/0207NHS).

### Sequencing

Sequencing was performed on either the *ONT MinION/GridION* or the *Illumina MiSeq* platform, as previously described. ^16^ Briefly, libraries were prepared in accordance with the ARTIC network protocols (v1 and v2) (https://artic.network/ncov-2019). For nanopore reads the ARTIC-nCov-2019 bioinformatics protocol was used, reads were size filtered, demultiplexed, trimmed with Porechop (https://github.com/rrwick/Porechop) and mapped against the reference strain Wuhan-Hu-1 (GenBank accession number MN908947), followed by clipping of primer regions. Variants were called using Nanopolish 0.11.3 (https://github.com/jts/nanopolish). For Illumina, reads were trimmed with trim_galore (bioinformatics.babraham.ac.uk/projects/trim_galore/) and mapped with BWA^20^ to the Wuhan-Hu-1 reference sequence, followed by primer trimming and consensus calling with iVar.^21^ A read coverage of least 10 was used for the consensus.

### Sequence Data

Consensus sequences with >90% coverage were included. All consensus genomes are available from the GISAID database (https://www.gisaid.org), the COG-UK consortium website (https://www.cogconsortium.uk/data/) and BAM files from the European Nucleotide Archive’s Sequence Read Archive service, BioProject PRJEB37886 (https://www.ebi.ac.uk/ena/data/view/PRJEB37886).

### Phylogenetic and probabilistic analysis

Retrospective phylogenetic analysis of whole-genome sequences was performed as follows. All full genomes of SARS-CoV-2 from Scotland sequenced as part of the COG-UK consortium were included and aligned using MAFFT v7.310 and with the HKY+I+G4 nucleotide substitution model determined by modeltest. The global lineage and UK lineage assignments for the dialysis samples were determined using civet (https://github.com/artic-network/civet).

We applied a novel algorithm, the Sequence Reporting Tool (SRT) to estimate the probability of healthcare-vs community-acquired infection in each case, based on the statistical approach developed for the COG-UK hospital-onset COVID-19 infection (HOCI) study (https://clinicaltrials.gov/ct2/show/NCT04405934).^22^ The approach is based on Bayesian principles and involves comparison of the proportion of similar viral sequences (with maximum pairwise SNP difference of two, with no difference where there is an overlap in ambiguous nucleotide codes or an ‘N’ in either sequence) observed within potential locations of infection for the case of interest: i) patients’ RDU and elsewhere in this hospital, ii) inpatient ward and hospital (if the patient was admitted) all within the prior three weeks; along with a weighted proportion of similar sequences in the local community of the patient within the prior six weeks based on the outer postcode of their home address. There are 61 districts based on this outcode (49 in Glasgow and 12 in Lanarkshire). We assumed 0.5 prior probability of infection within the RDU before consideration of sequence data, and the prior probability of admission-related infection among the inpatients was based on the interval from admission to diagnosis and the incubation distribution of COVID-19.^23^ The SRT algorithm outputs two posterior probabilities in all cases: that of acquiring the virus directly from the RDU (p_RDU) and that of acquiring the infection through use of facilities within the hospital but *not* in the RDU (e.g. toilets, cafes, lobbies, or shared transport, etc) (p_hRDU). A posterior probability (p_RDU) of 1 indicates that the transmission occurred within the RDU; if the p_RDU stays at 0.5 then it remains unclear where the transmission occurred; if p_RDU is 0 then the infection was most likely community acquired. If the haemodialysis patient was an inpatient and continued to attend the RDU at the time of COVID-19 diagnosis, the SRT algorithm also gives the probability of acquiring the infection from the ward of admission (p_wADM) and that of infection elsewhere in that hospital (p_hADM). The SRT algorithm was coded in R version 3.6.0, using the ape v5.3 package for calculation of pairwise SNP differences and PostcodesioR v0.1.1 and gmt v2.0-1 packages to calculate distances between postcodes.^22^

### Survival statistics

Comparisons were made between patients who lived and died following SARS-CoV-2 infection. At the time of analysis, no patients who were still alive were critically ill or requiring oxygen therapy. The mortality rates were calculated for patients requiring dialysis expressed as deaths per 1000 patient days were calculated over the three months 1^st^ March-31^st^ May 2020.

## Results

### Description of cases, treatments and outcomes in patients requiring haemodialysis with COVID-19

In total, 60 of 671 (8.9%) patients requiring HD were diagnosed with COVID-19 infection during 1^st^ March-31^st^ May 2020. 16/60 patients (26.7%) died; with COVID-19 as the certified cause of death. There were no statistically significant differences in the clinical characteristics and associated co-morbidities between those who died and those who survived (Table 1). The median time from positive SARS-CoV-2 test to death was 10.5 days (range 0-29 days). Two patients required intensive care (of whom one died). No patients received ‘specific’ therapy for COVID-19 (e.g. dexamethasone, remdesivir, tocilizumab). Compared to the corresponding three-month periods 2018-19 (mean deaths 44/quarter year), there were 16 more deaths in patients undergoing outpatient haemodialysis in the same RDUs, equivalent to 0.797 deaths per 1000 patient days in all patients requiring haemodialysis during 1^st^ March-31^st^ May 2020 compared to 0.628 deaths per 1000 patient days as mean of the corresponding period during 2017-2018 (equivalent to a 27.0% increase).

**Table 1.**
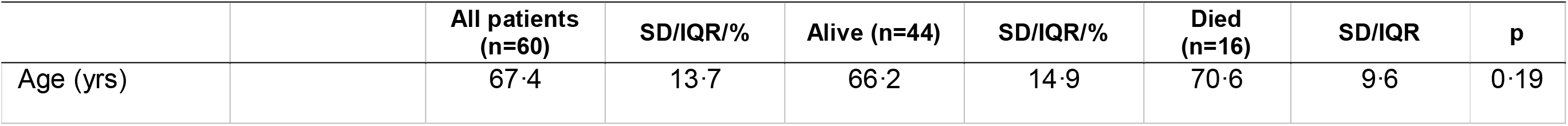
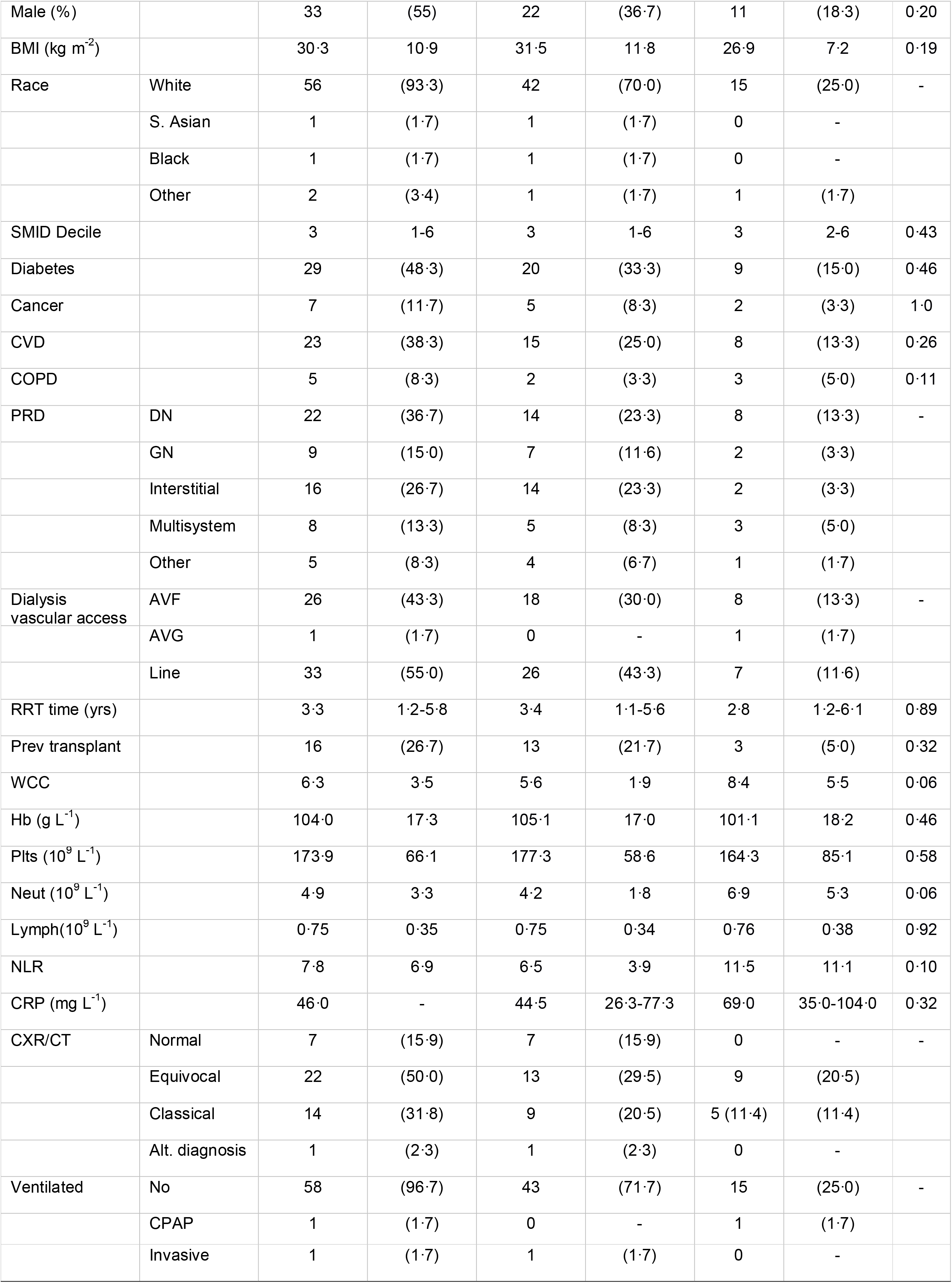
Demographic, laboratory and imaging data of patients with COVID-19 with comparisons between patients who died compared to survivors. Laboratory test results are taken from the date closest to diagnosis of SARS-CoV2 (same day in 85% of cases with only two cases where laboratory day >2 days from date of diagnosis). All patients requiring haemodialysis are registered on the Strathclyde Electronic Renal Patient Record (SERPR; Vitalpulse, UK) which records clinical, laboratory and imaging data for clinical care, audit and research. Using SERPR we extracted anonymised clinical data on all patients treated with haemodialysis with a positive test for SARS-CoV-2 infection. The Scottish Government provides online calculators allowing use of patient postcode to generate divisions of socioeconomic deprivation, the Scottish Index of Multiple Deprivation (SIMD) (http://www.gov.scot/Topics/Statistics/SIMD). Deprivation deciles were calculated and categorized into most deprived deciles with 1 corresponding to most deprived and 10 as least deprived. Cause of death was certified by each patient’s clinical team with registration with the Scottish Mortality Audit in Renal Replacement Therapy (SMARRT).^30^ Radiological imaging was coded using the British Thoracic Society Classification for COVID-19. (https://www.bsti.org.uk/media/resources/files/BSTI_COVID_CXR_Proforma_v.3-1.pdf). Values are presented as means (standard deviation) or medians (inter-quartile range) and comparisons between groups were made using t-test, Kruskal-Wallis, Chi-square and Fisher’s exact test as appropriate. Statistics were performed on Minitab Version 19.2020.1.0 (Minitab, State College, Pennsylvania). Abbreviations BMI – body mass index, CVD - cardiovascular disease, COPD - chronic obstructive pulmonary disease, PRD - primary renal disease, DN – diabetic nephropathy, GN - glomerulonephritis, AVF - arteriovenous fistula, AVG - arteriovenous graft, RRT - renal replacement therapy, WCC – white cell count, Hb – haemoglobin, Plts - platelets, Neut – neutrophils, Lymph – lymphocytes, NLR – neutrophil to lymphocyte ratio, CXR - chest X-ray, CT –computed tomography of chest, CPAP – continuous positive airway pressure

**Figure 3**
|  |  | All patients<br>(n=60) | SD/IQR/% | Alive (n=44) | SD/IQR/% | Died<br>(n=16) | SD/IQR | p |
| --- | --- | --- | --- | --- | --- | --- | --- | --- |
| Age (yrs) |  | 67.4 | 13.7 | 66.2 | 14.9 | 70.6 | 9.6 | 0.19 |

Table 1
|  |  |  |  |  |  |  |  |  |
| --- | --- | --- | --- | --- | --- | --- | --- | --- |
| Male (%) |  | 33 | (55) | 22 | (36.7) | 11 | (18.3) | 0.20 |
| BMI (kg m<sup>-2</sup>) |  | 30.3 | 10.9 | 31.5 | 11.8 | 26.9 | 7.2 | 0.19 |
| Race | White | 56 | (93.3) | 42 | (70.0) | 15 | (25.0) | - |
|  | S. Asian | 1 | (1.7) | 1 | (1.7) | 0 | - |  |
|  | Black | 1 | (1.7) | 1 | (1.7) | 0 | - |  |
|  | Other | 2 | (3.4) | 1 | (1.7) | 1 | (1.7) |  |
| SMID Decile |  | 3 | 1-6 | 3 | 1-6 | 3 | 2-6 | 0.43 |
| Diabetes |  | 29 | (48.3) | 20 | (33.3) | 9 | (15.0) | 0.46 |
| Cancer |  | 7 | (11.7) | 5 | (8.3) | 2 | (3.3) | 1.0 |
| CVD |  | 23 | (38.3) | 15 | (25.0) | 8 | (13.3) | 0.26 |
| COPD |  | 5 | (8.3) | 2 | (3.3) | 3 | (5.0) | 0.11 |
| PRD | DN | 22 | (36.7) | 14 | (23.3) | 8 | (13.3) | - |
|  | GN | 9 | (15.0) | 7 | (11.6) | 2 | (3.3) |  |
|  | Interstitial | 16 | (26.7) | 14 | (23.3) | 2 | (3.3) |  |
|  | Multisystem | 8 | (13.3) | 5 | (8.3) | 3 | (5.0) |  |
|  | Other | 5 | (8.3) | 4 | (6.7) | 1 | (1.7) |  |
| Dialysis vascular access | AVF | 26 | (43.3) | 18 | (30.0) | 8 | (13.3) | - |
|  | AVG | 1 | (1.7) | 0 | - | 1 | (1.7) |  |
|  | Line | 33 | (55.0) | 26 | (43.3) | 7 | (11.6) |  |
| RRT time (yrs) |  | 3.3 | 1.2-5.8 | 3.4 | 1.1-5.6 | 2.8 | 1.2-6.1 | 0.89 |
| Prev transplant |  | 16 | (26.7) | 13 | (21.7) | 3 | (5.0) | 0.32 |
| WCC |  | 6.3 | 3.5 | 5.6 | 1.9 | 8.4 | 5.5 | 0.06 |
| Hb (g L<sup>-1</sup>) |  | 104.0 | 17.3 | 105.1 | 17.0 | 101.1 | 18.2 | 0.46 |
| Plts (10<sup>9</sup> L<sup>-1</sup>) |  | 173.9 | 66.1 | 177.3 | 58.6 | 164.3 | 85.1 | 0.58 |
| Neut (10<sup>9</sup> L<sup>-1</sup>) |  | 4.9 | 3.3 | 4.2 | 1.8 | 6.9 | 5.3 | 0.06 |
| Lymph(10<sup>9</sup> L<sup>-1</sup>) |  | 0.75 | 0.35 | 0.75 | 0.34 | 0.76 | 0.38 | 0.92 |
| NLR |  | 7.8 | 6.9 | 6.5 | 3.9 | 11.5 | 11.1 | 0.10 |
| CRP (mg L<sup>-1</sup>) |  | 46.0 | - | 44.5 | 26.3-77.3 | 69.0 | 35.0-104.0 | 0.32 |
| CXR/CT | Normal | 7 | (15.9) | 7 | (15.9) | 0 | - | - |
|  | Equivocal | 22 | (50.0) | 13 | (29.5) | 9 | (20.5) |  |
|  | Classical | 14 | (31.8) | 9 | (20.5) | 5 (11.4) | (11.4) |  |
|  | Alt. diagnosis | 1 | (2.3) | 1 | (2.3) | 0 | - |  |
| Ventilated | No | 58 | (96.7) | 43 | (71.7) | 15 | (25.0) | - |
|  | CPAP | 1 | (1.7) | 0 | - | 1 | (1.7) |  |
|  | Invasive | 1 | (1.7) | 1 | (1.7) | 0 | - |  |

### Genomic and epidemiological investigation

Residual RNA extract from 53 of 60 patients with SARS-CoV-2 positive samples were obtained for virus genome sequencing. 39 of these sequences plus one from a healthcare worker were of sufficient quality and coverage for further analysis. The samples belonged to 13 different UK lineages (Figure 2). Whilst a number of patients had indistinguishable sequences from the same UK lineage and shared dialysis sessions, some fell outwith phylogenetic lineages, providing evidence of community-transmission. The recent introduction of SARS-CoV-2 into the human population and its relatively low mutation rate, mean sequences in the same UK lineage and phylotype cannot be interpreted as direct transmission events, with further temporal and epidemiological data required to quantify the probability of transmission. Conversely, sequences from different UK lineages would disprove direct transmission. Epidemiological investigation identified clusters of SARS-CoV-2 positive patients with shared dialysis sessions and sometimes transport and this was analysed with the phylogenetic data (Figure 2 and 3). We found five of the six RDUs spanning two health boards in the West of Scotland had evidence of unit-linked transmission events.

**Figure 1.**
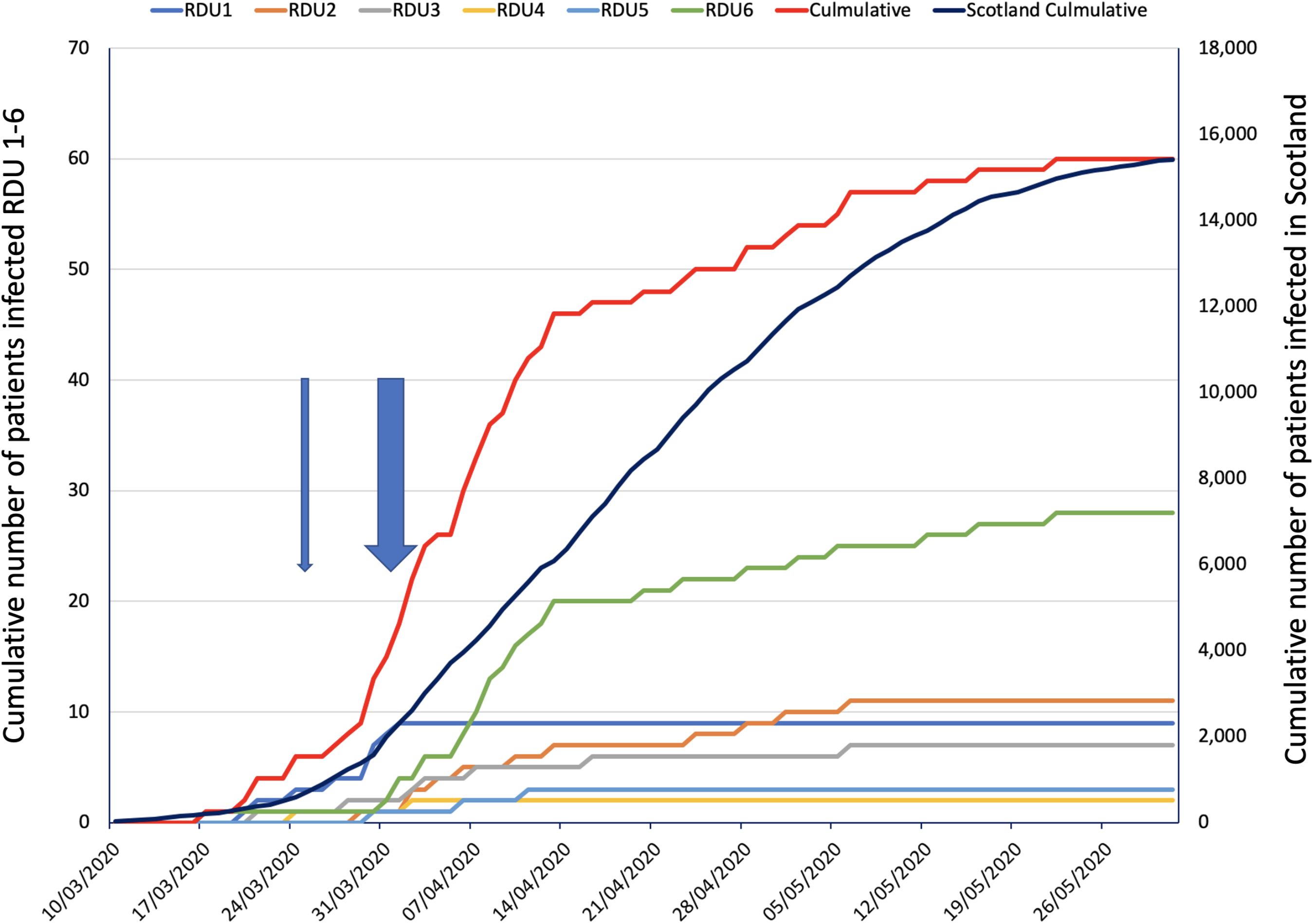
Cumulative cases of COVID-19 cases (left y-axis) with arrows demonstrating additional infection control measures (narrow arrow - RDU1, wide arrow covers the dates for all other RDUs). Cumulative infection numbers for Scotland are on the right y-axis.

**Figure 2.**
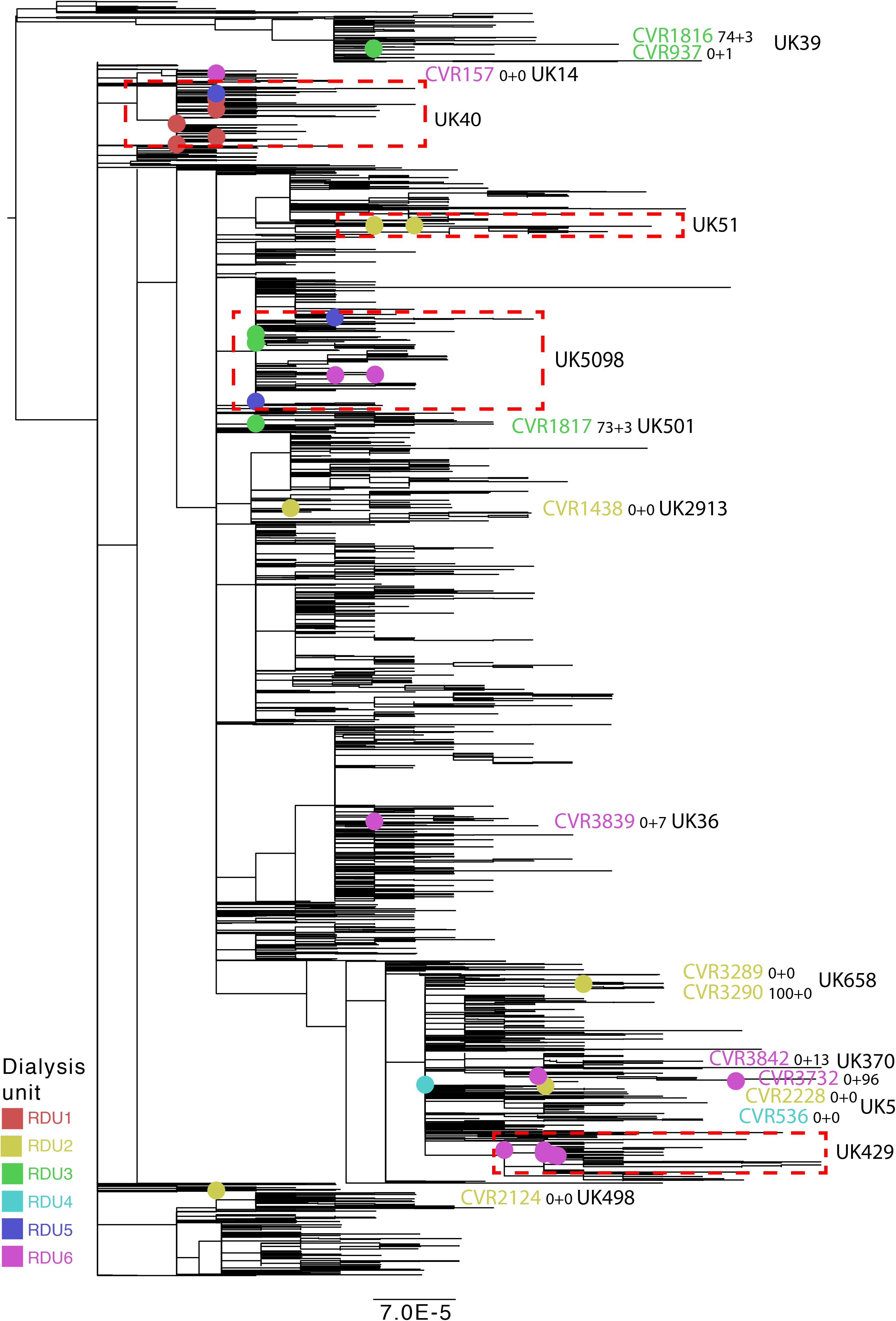
Phylogenetic tree showing the relationship of 39 sequences from RDU patients and additional SARS-CoV-2 genomes from Scotland. Sequences are colour-coded by RDU location. Dashed boxes highlight the UK lineage and are shown in more detail in Figure 3. The numerical suffixes of the CVR identifier indicate the posterior probability (as a percentage) of the patient acquiring SARS-CoV-2 from the RDU (p_RDU) or from the wider hospital where dialysis takes place (p_hRDU). The scale bar indicates substitutions per nucleotide site.

**Figure 3.**
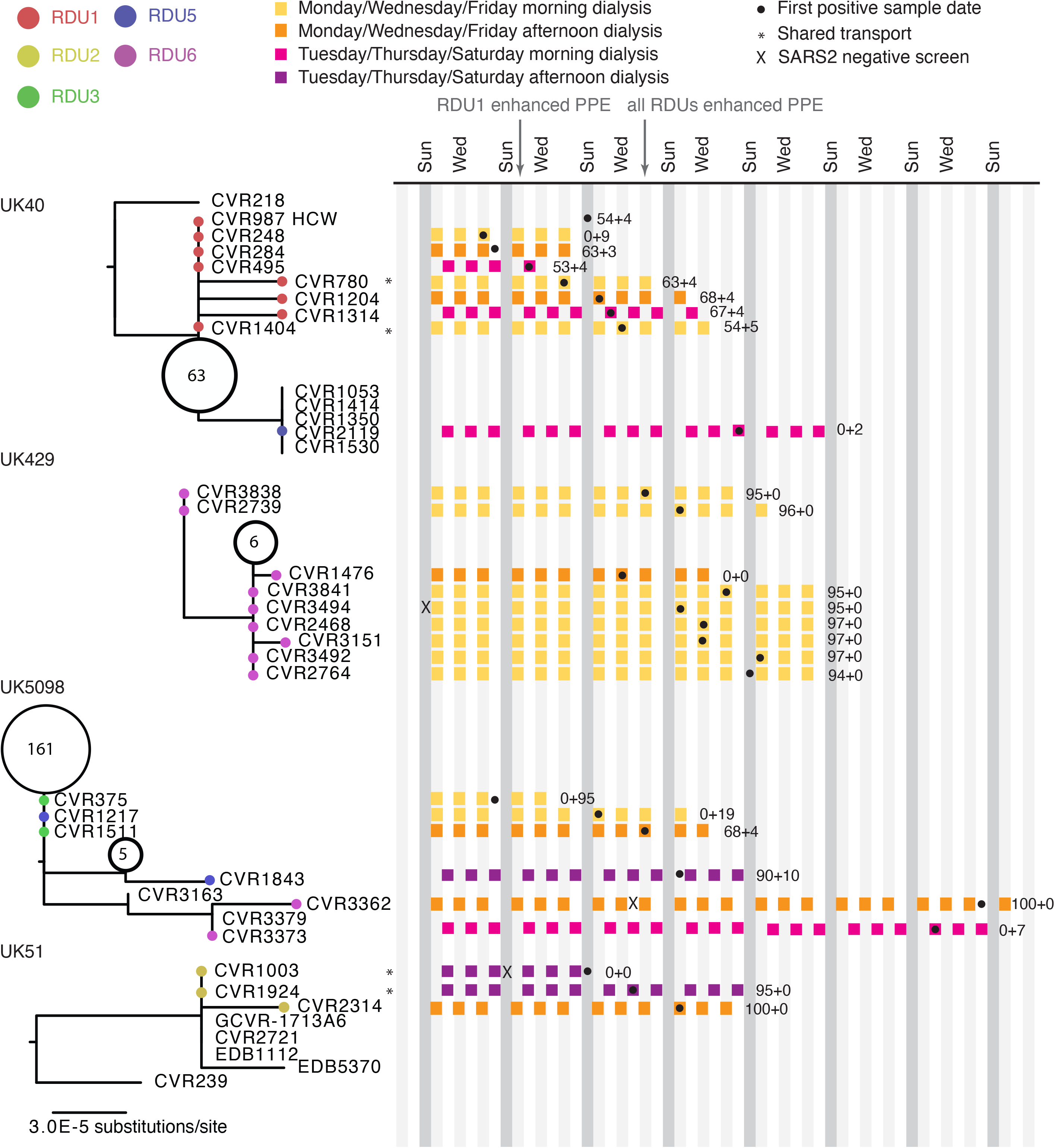
Timeline of detection of first SARS-CoV-2 positive results in haemodialysis patients in RDUs with details of dialysis sessions and shared patient transport in relation to the UK lineage. The phylogenetic trees are derived from the dashed boxes in Figure 2. Circled numbers in the phylogenetic tree represent the number of indistinguishable sequences from Scotland for the given node on the phylogeny. The numerical suffixes of the CVR identifier indicate the posterior probability (as a percentage) of the patient acquiring SARS-CoV2 from the RDU or from another healthcare-related infection (i.e., hospital where dialysis takes place and ward and/or hospital they have been admitted to), respectively. The scale bar indicates substitutions per nucleotide site.

### RDU1

In RDU1, viral sequences from seven haemodialysis patients and one HCW from the same unit clustered within the UK40 lineage (Figure 3). Five of these sequences were indistinguishable to each other (CVR248, CVR284, CVR495, CVR987 and CVR1404), suggestive of within-unit transmission. Applying the SRT, we found the probability of within-unit transmission in RDU1 was indeterminate based on sequence data alone ranging from 0.53 to 0.68. Further epidemiological analysis suggested transmission in some cases – for example, CVR248 and CVR1404 had indistinguishable sequences and shared dialysis sessions (Figure 3). However, CVR284 and CVR495, also with indistinguishable sequences, did not overlap with each other or anyone else on the unit, suggesting community-acquisition. HCW, CVR987, having been in direct contact with CVR248, self-isolated on 22^nd^ March, prior to testing positive seven days later. However, this viral sublineage of UK40, was widespread in the community, with 63 other indistinguishable sequences detected within the geographical location of RDU1 and patient communities; so this transmission could not reliably be inferred. Two of the seven patients whose sequences derived from the wider UK40 lineage (CVR780 and CVR1404) (Figure 3) were linked epidemiologically, sharing both dialysis sessions and transport from home to the unit. However, the estimated probabilities of within unit transmission for CVR780 and CVR1404 were 0.63 and 0.54, respectively. This result was supported by close inspection of the data. CVR780 was found to have tested positive for SARS-CoV-2 six days *prior* to CVR1404 and had a single nucleotide polymorphism relative to the Wuhan reference not found in CVR1404, making transmission from CVR780 to CVR1404 less likely. The widespread distribution of UK40 lineages in the community, the early preventative measures implemented by RDU1 and the lack of definitive epidemiological evidence for transmission suggest that individual transmissions were not due to infection prevention and control (IPC) challenges in this unit. The final case occurred nine days after “lockdown” and the implementation of enhanced PPE measures.

### RDU2

In RDU2, there was evidence of five introductions of SARS-CoV-2 from the community, of which two lineages spread within the unit or on hospital transport to the unit. CVR3289 and CVR3290 had indistinguishable sequences, only seen in two other non-geographically linked community cases. These patients shared the same dialysis session and transport, with an estimated 100% probability of within-unit transmission (Figure 2, lineage UK658). Likewise, CVR1003 and CVR1924 had indistinguishable sequences and shared the same dialysis sessions (Figure 3, lineage UK51). CVR2314 had a p_RDU of 1, but this patient had no epidemiological link to other two patients, suggesting nosocomial transmission from fomites or an untested staff member (staff were not routinely tested for SARS-CoV-2, at this time).

### RDU3, 4 and 5

In RDU3, three introductions and two separate transmission events were identified. Although CVR375 and CVR1511 had indistinguishable sequences, this was shared with 161 other Scottish samples (Figure 2, lineage UK5098). The SRT identified nosocomial infection in CVR375 due to within-hospital rather than within-dialysis unit transmission (p_hADM 0.95). CVR1511 acquisition of infection from RDU3 (p_RDU 0.68) was less clear. CVR1817 is a close sequence match to CVR375 and CVR1511 based on the 2 SNP threshold, leading the SRT to estimate probable unit-based transmission (p_RDU = 0.73). However, on phylogenetic analysis CVR1817 falls into the separate UK501 lineage (Figure 2) and appears likely to have been community-acquired on consideration of all available information. In lineage UK39, CVR937 was community-acquired while the related CVR1816 was probable within-unit transmission (p_RDU = 0.74).

In RDU4, two patients tested positive for SARS-CoV-2 but there was no linkage found on sequence analysis (Supplementary Table 1) with no evidence of within unit transmission of SARS-CoV-2 in this dialysis unit.

Within RDU5, three cases of SARS-CoV-2 infection were detected. CVR1217 (lineage UK5098) was community-acquired while CVR1843 (p_RDU of 0.9) was highly suggestive of within RDU transmission. These related sequences differed by 2 SNPs. This strongly suggests that intermediary modes of transmission should ideally have been investigated, including untested asymptomatic individuals, for example members of staff.

### RDU6

SARS-CoV-2 was introduced to RDU6 on at least 5 occasions with evidence of onward transmission in two cases and hospital-transmission in one. Of the 26 (12°3% of total) SARS-CoV-2 positive patients, 16 were sequenced. Nine of these patients were within the UK429 lineage (Figure 3). The SRT verified a high likelihood of within-unit transmission, with p_RDU ranging from 0°96 to 1 (Figure 3). CVR3373 (p_RDU = 0, the first of this phylotype found in RDU6) and CVR3362 were within a separate lineage, UK5098 (Figure 3). CVR3362 had been hospitalised for a month prior to the positive SARS-CoV-2 test, whilst maintaining dialysis within RDU6 and had a p_RDU of 1. Further discussion with the infection control team confirmed that CVR3379 (non-haemodialysis patient) and another unsequenced SARS-CoV-2 positive haemodialysis patient, shared the same hospital bay with CVR3362. It is possible that SARS-CoV-2 was brought onto this bay by the other haemodialysis patient from their dialysis sessions on RDU6. This highlights one of the limitations of phylogeny and the SRT algorithm in an outbreak investigation – sequencing data needs to be representative of prevalent cases for the results to be interpreted. Finally, CVR3732 (Figure 2, UK370) had a high p_hRDU (0°96) indicating acquisition of SARS-CoV-2 from elsewhere in the hospital.

### Summary

We found evidence of multiple introductions of SARS-CoV-2 infection into Scottish dialysis units and of onward transmission within these units. There was strong evidence for 15 patients acquiring SARS-CoV-2 in hospital or on shared hospital transport. For a further 9 patients the source of infection was less certain although most likely acquired within the hospital RDU setting (In lineage UK40: CVR284, CVR495, CVR780, CVR1204, CVR1314, CVR1404 were similar but multiple indistinguishable sequences were also detected in the community; lineage UK5098: CVR1511; lineage UK39: CVR1816; lineage UK501: CVR1817). A further 15 patients most likely acquired SARS-CoV-2 in the community (Supplementary Table 1). RDU6 cases had a high likelihood of within-unit transmission of SARS-CoV-2 and RDU6 also had one of the highest rates of infection over the longest time period (31^st^ March to 26^th^ May, Table 2). However, in RDU1, where the rate of infection was also high, there was tentative evidence of within-unit transmission; infections occurred over 12 days, the incubation period of the last case was coincident with both “lockdown” and enhanced PPE implementation.

**Table 2.**
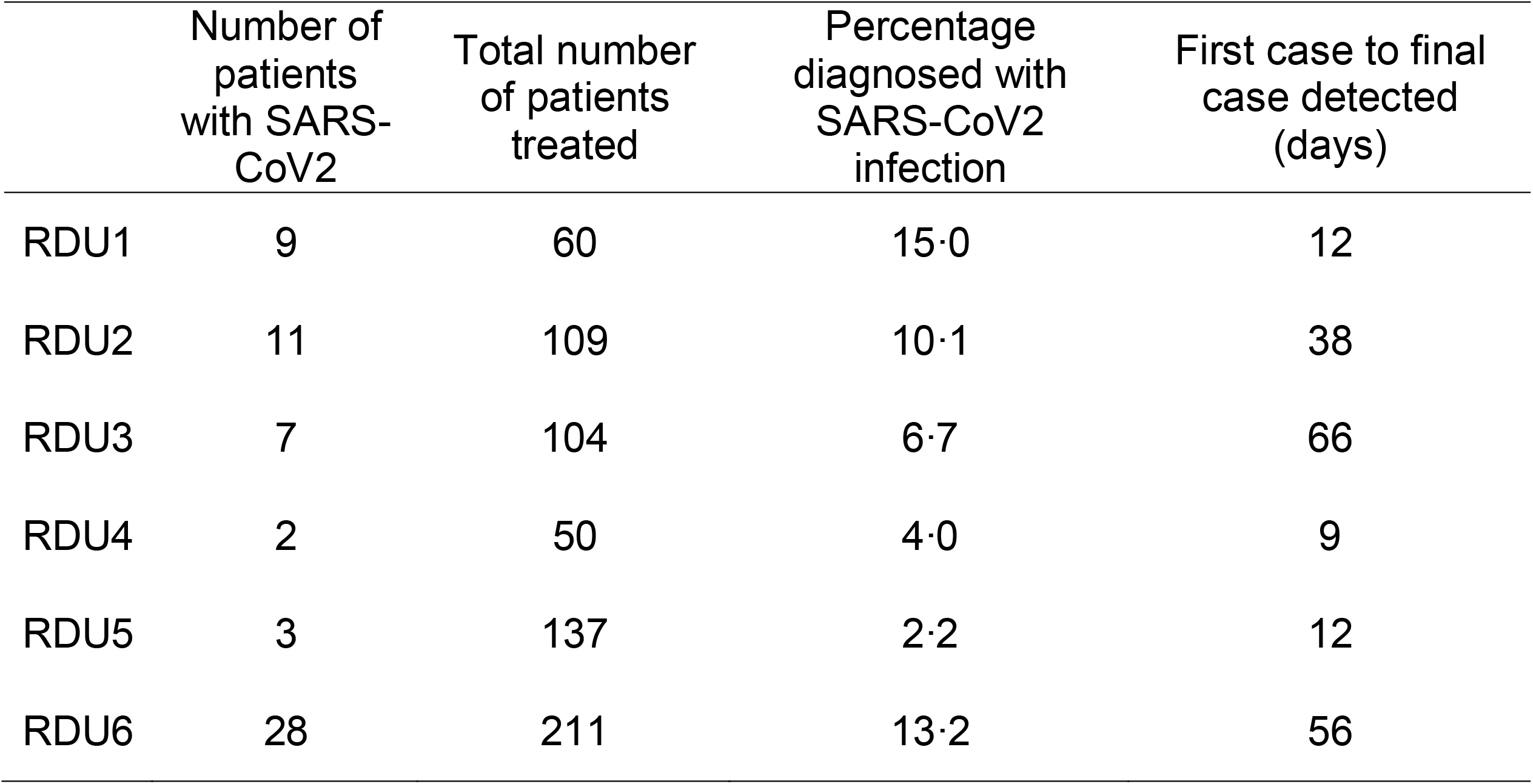
Number of patients treated at each RDU and proportion of patients infected with SARS-CoV-2 per RDU.

## Discussion

During the first wave of the UK pandemic, we studied SARS-CoV-2 infections within 6 affected Scottish RDUs. Using a genomic epidemiology approach, we found that transmission of SARS-CoV-2 within RDUs was common, affecting 8°9% of dialysis patients with a very high associated mortality (27%) in keeping with other recent studies. ^8-11^Many guidelines have evolved for the care of dialysis patients to minimise risk of infection, including nosocomial transmission of SARS-CoV-2.^5,6,24^ Less is known around whether patients requiring dialysis are at greater risk of community transmission of SARS-CoV-2 and how infection in the dialysis units relates to viral exposure in the healthcare environment compared to that in the community. Whole-genome sequencing provides high-level resolution of SARS-CoV-2 genome and in combination with epidemiological data can facilitate our understanding of transmission and evolution during pathogen outbreaks.^13,14^ Applying careful analysis of data generated from the community and the hospital setting, we found risk was present both from community and hospital settings; almost always from within the RDUs themselves, but on occasion from the wider hospital. Multiple introductions occurred into the dialysis units, reflecting the risks associated with individuals having increased contact with the community (including need for frequent travel to hospitals) as well as prolonged and regular contact within the RDUs themselves. Data from the Scottish Renal Registry suggest that measures introduced in early April to reduce this exposure, including the use of masks to and from dialysis and individual transport, were effective; with the number of cases in people receiving dialysis falling sharply two to three weeks before the rest of the general population within Scotland.^25^ In order to capitalise on the utility of such knowledge in hospital-outbreak management, the results from the sequencing data need to be generated in a timely manner.^15,17^ However, the feasibility of implementing this rapid sequencing response in the National Health Service to date has been impeded by lack of expertise and equipment for both high-throughput whole-genome sequencing and for processing the data generated into a form interpretable by infection control and public health. COG-UK has demonstrated that near real-time sequencing is achievable at scale.^17^ We used this framework and a novel statistical algorithm to characterise transmission dynamics specifically in the haemodialysis cohort, a group both at higher risk of severe outcome as well as having numerous healthcare interactions.

A limitation of the study is the sequences available for analysis; accurate estimation of the likely source of infection depends on having sufficient sequences available from the community of affected patients and of cases from the hospital setting. Here, we obtained sequences from two-thirds of the lab-detected SARS-CoV-2 cases from RDU1-RDU5, and 44% for RDU6. We also compared data from the RDUs with 944 other cases in the community, 700 inpatients and 546 samples taken from patients presenting in emergency departments in the same health boards as provide care for dialysis patients. Additionally, COVID-19 is asymptomatic in up to 20% of patients, which may have reduced the number of infections captured.^26^ Further, early in the pandemic, HCWs were ineligible for testing. Frequent, regular testing of all HCWs and all patients, regardless of symptoms is warranted. There is also mounting evidence that HCWs have a higher seroprevalence for SARS-CoV-2 antibodies than the general population, with a high proportion being asymptomatic.^27,28^

The low substitution rate of SARS-CoV-2 limits the granularity of outbreak analysis; as demonstrated in this study, indistinguishable sequences may not be part of a transmission cluster if there is widespread circulation in the patients’ home communities. To address this limitation, we employed the SRT, which combines sequence data with both temporal and geographic data to improve estimates of within-unit and within-hospital versus community transmission. Based on additional epidemiological evidence such as timing of haemodialysis sessions and hospital transport, the SRT correctly identified a high probability of within-unit transmission for RDU 2, 3, 5 and 6. Less definitive results for RDU1, not immediately apparent by phylogenetic investigation alone (due to the widespread presence the lineage within the community), affirm its potential as a rapid tool to aid outbreak investigations. We confirm the findings of other published reports of SARS-CoV-2 in the haemodialysis cohort that cases are at risk of poor outcomes, with no specific at-risk group identified based on comorbid conditions.^9^ The high mortality, dearth of therapeutics and likely poor response to vaccines,^29^ emphasises the need for targeted strategies to mitigate risk in this cohort. Identification of major transmission risks is vital to address outbreaks in this vulnerable group where there is prolonged, unavoidable contact between healthcare settings and the local community. Whilst universal infection control measures are beneficial, we identified multiple community-acquired infections, with RDUs being an interface for transmission. Additional measures may be required to reduce infection in this setting. Longer, more extreme periods of intensive social distancing (‘shielding’) to reduce contact with other individuals may be required when the community incidence of infection is high. Knowledge of the dominant site of transmission can justify and provide precision to the recommendation to shield and condense the period of isolation, loneliness and distress in this cohort, who already have a high incidence of depression.

Although we demonstrate the utility of identifying the likelihood of transmission of SARS-CoV-2 infection around treatment centres for haemodialysis, our findings have resonance for any group requiring frequent attendance in healthcare facilities, such as patients undergoing chemotherapy, radiotherapy or outpatient rehabilitation. (Word count: 3484)

## Supporting information

STROBE_CHECKLIST

COGUK_authorship

## Data Availability

Data sharing statement Will individual participant data be available? Yes What data in particular will be shared? Individual participant data that underlie the results reported in this article, after de-identification. Sequences and de-identified metadata is available on MRC-CLIMB through COG-UK. Sequences are also available on GISAID. What other documents will be available? Study protocol, statistical analysis, analytic code When will data be available? Immediately following publication, no end date With whom? Researchers who provide a methodologically sound proposal For what types of analysis? To achieve aims in the approved proposal By what mechanism will data be made available? Proposals should be directed to (Statement from COG-UK consortium: We are committed to open science, and sharing all data that we can as rapidly as possible. This includes sharing data for use by Public Health authorities internationally, to support COVID-19 response, and sharing data in such a way that the academic community can access and use the data and analysis according to FAIR data principles.)

https://gla-my.sharepoint.com/:w:/g/personal/kathy_li_glasgow_ac_uk/EUKpswq8Hq9Atpd2xO7FHusByY1AP3vUFrUQy11yhqn79Q

https://gla-my.sharepoint.com/:x:/g/personal/kathy_li_glasgow_ac_uk/EZ-b-VbxynZEjLiF4uy82FsB__39HZJmKiRJrC-zh1QMXg

## Author contributions

Each named author has substantially contributed to conducting the underlying research and drafting this manuscript. Additionally, to the best of our knowledge, the named authors have no conflict of interest, financial or otherwise.

## Ethical approval

Use of anonymised data was approved by the Local Privacy Advisory Group of NHS Greater Glasgow and Clyde ‘Safe Haven’ on behalf of the West of Scotland Ethics Committee (approval GSH/20/RE/001). Surplus RNA extract was collected for sequencing with ethical approval from the NHSGGC biorepository (16/WS/0207NHS).

## Funding

Funding: COG-UK is supported by funding from the Medical Research Council (MRC) part of UK Research & Innovation (UKRI), the National Institute of Health Research (NIHR) and Genome Research Limited, operating as the Wellcome Sanger Institute. Also MRC (MC UU 1201412), UKRI/Wellcome (COG-UK), Wellcome Trust Collaborator Award (206298/Z/17/Z – ARTIC Network; TCW Wellcome Trust Award 204802/Z/16/Z.

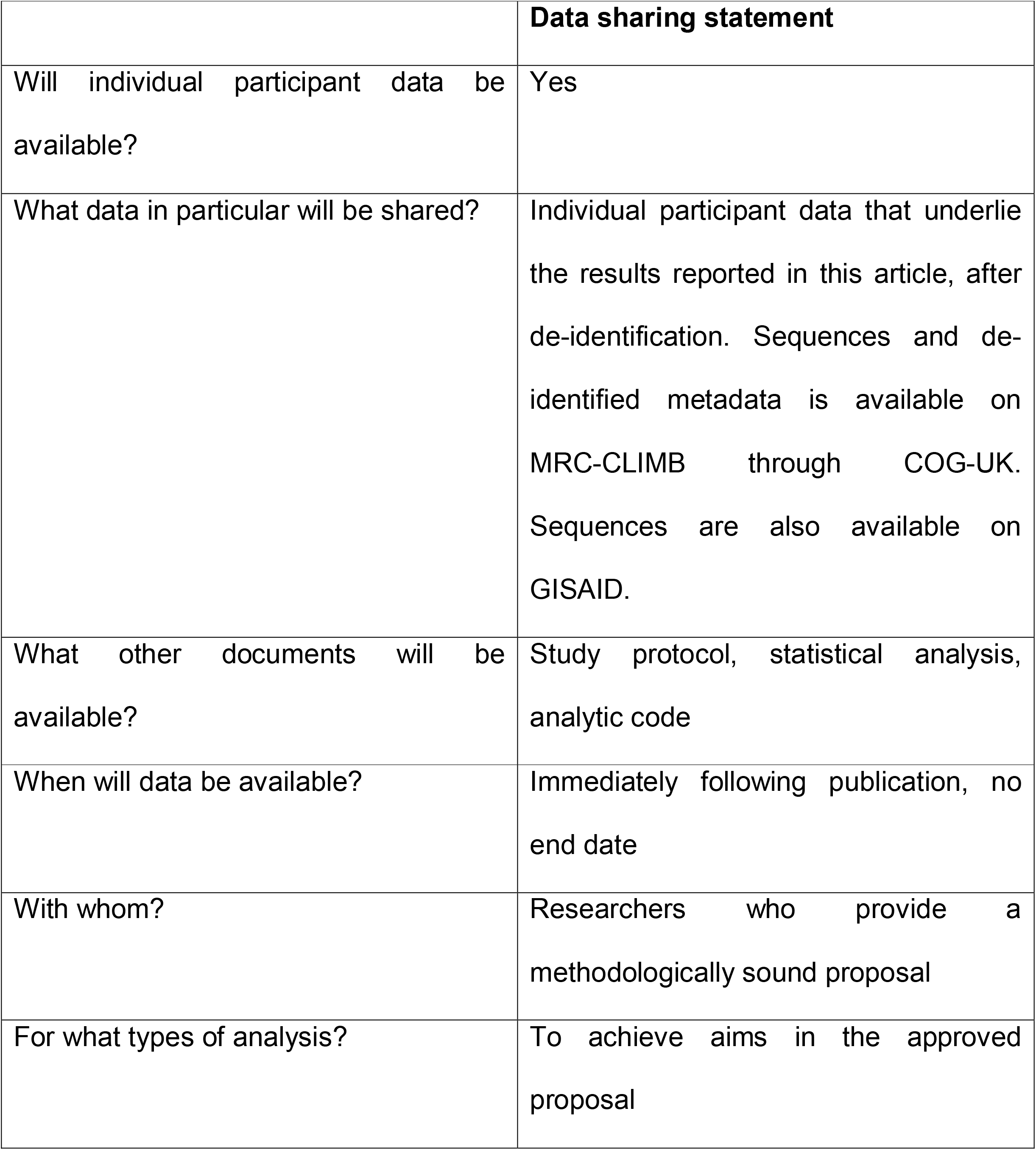
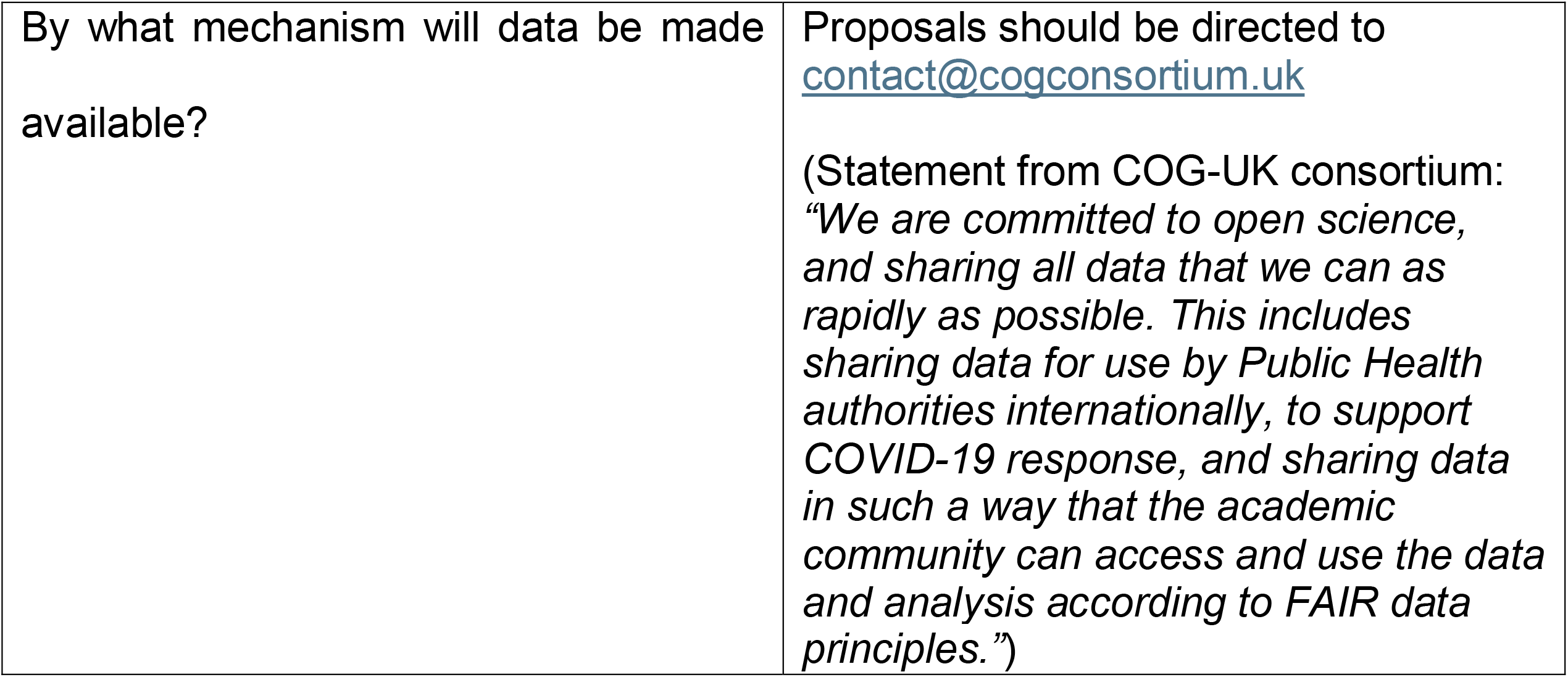

## Footnotes

i MagNA Pure 96 system (Roche, Penzberg, Germany), Abbott M2000 (Abbott, Chicago, US) or Cobas^®^ 6800 Systems (Roche). SARS-CoV-2 was detected using one of three RT-PCR assays: RdRp gene/E gene.^19^ RdRp gene/N gene (Abbott RealTime SARS-CoV-2 assay, Chicago, US) or the ORF1a/b and E gene (Cobas^®^ SARS-CoV-2, Roche).

